# Grounded large language models for diagnostic prediction in real-world emergency department settings

**DOI:** 10.1101/2025.02.23.25322736

**Authors:** Alexandre Niset, Ines Melot, Margaux Pireau, Alexandre Englebert, Nathan Scius, Julien Flament, Salim El Hadwe, Mejdeddine Al Barajraji, Henri Thonon, Sami Barrit

**Affiliations:** Emergency Medicine, Université Catholique de Louvain, Belgium; Pediatric Intensive Care Unit, Cliniques Universitaires Saint-Luc, Brussels, Belgium; Sciense, New-York; Emergency Department, CHU Helora – Jolimont, La Louvière, Belgium; Emergency Department, Cliniques Universitaires Saint-Luc, Brussels, Belgium; Information and Communication Technologies, Electronics and Applied Mathematics (ICTEAM), UCLouvain, Louvain-la-Neuve, Belgium; Orthopedic department, CHU UCL Namur, Yvoir, Belgium; Emergency Department, CHU UCL Namur, Yvoir, Belgium; Department of Clinical Neurosciences, University of Cambridge, Cambridge, CB2 1TN, UK; Department of Neurosurgery, CHR Citadelle, Liege, Belgium; Neurosurgery, Université Libre de Bruxelles, Brussels, Belgium

## Abstract

**Background:** Emergency departments face increasing pressures from staff shortages, patient surges, and administrative burdens. While large language models (LLMs) show promise in clinical support, their deployment in emergency medicine presents technical and regulatory challenges. Previous studies often relied on simplistic evaluations using public datasets, overlooking real-world complexities and data privacy concerns.

**Methods:** At a tertiary emergency department, we retrieved 79 consecutive cases during a peak 24-hour period constituting a siloed dataset. We evaluated six pipelines combining open- and closed-source embedding models (text-embedding-ada-002 and MXBAI) with foundational models (GPT-4, Llama3, and Qwen2), grounded through retrieval-augmented generation with emergency medicine textbooks. The models’ top-five diagnostic predictions on early clinical data were compared against reference diagnoses established through expert consensus based on complete clinical data. Outcomes included diagnostic inclusion rate, ranking performance, and citation sourcing capabilities.

**Results:** All pipelines showed comparable diagnostic inclusion rates (62.03-72.15%) without significant differences in pairwise comparisons. Case characteristics, rather than model combinations, significantly influenced predictive diagnostic performance. Cases with specific diagnoses were significantly more diagnosed versus unspecific ones (85.53% vs. 31.41%, p<0.001), as did surgical versus medical cases (79.49% vs. 56.25%, p<0.001). Open-source foundational models demonstrated superior sourcing capabilities compared to GPT-4-based combinations (OR: 33.92 to ∞, p<1.4e-12), with MBXAI/Qwen2 achieving perfect sourcing.

**Conclusion:** Open and closed-source LLMs showed promising and comparable predictive diagnostic performance in a real-world emergency setting when evaluated on siloed data. Case characteristics emerged as the primary determinant of performance, suggesting that current limitations reflect AI alignment fundamental challenges in medical reasoning rather than model-specific constraints. Open-source models’ demonstrated superior sourcing capabilities—a critical advantage for interpretability. Continued research exploring larger-scale, multi-centric efforts, including real-time applications and human-computer interactions, as well as real-world clinical benchmarking and sourcing verification, will be key to delineating the full potential of grounded LLM-driven diagnostic assistance in emergency medicine.

## Background

Since the early 2000s, emergency medicine has faced a persistent crisis exacerbated by the COVID-19 pandemic ^1–4^. Emergency departments are challenged by staff shortages and surges in patient loads, all while intensifying technical demands and administrative burdens^2^. In these resource-constrained, high-paced environments, emergency physicians (EPs) must make critical, time-sensitive decisions across primary and acute care contexts^5^.

Amid these pressures, AI has shown promise as a supportive tool^6^. Notably, large language models (LLM) with advanced natural language processing capabilities may bolster clinical decision-making and streamline communication^7–9^. Nevertheless, deploying LLM in specialized fields such as EM presents critical challenges due to stringent technical, ethical, and regulatory constraints to meet exacting domain-specific requirements^10^.

In this context, the commercialization of frontier models has made state-of-the-art AI technologies widely accessible^11^ and prompted many biomedical investigators to hastily leverage these proprietary solutions in their research^12,13^. In turn, many studies rely on simplistic paradigms using convenient benchmarks, such as public question banks. These often overlook real-world complexities and expose them to data contamination, overfitting, and self-fulfilling prophecies^14,15^.

In parallel, regulatory frameworks increasingly mandate AI interpretability^16^ while also governing personal data confidentiality, particularly regarding data sharing with private entities or across borders^17,18^. Commercial solutions may exploit user interactions obscurely for undisclosed objectives—from model training to business intelligence^19^—risking the exposure of sensitive data, especially with naive users^20,21^.

While large-scale efforts offer significant promise, they are predominantly driven by major technological firms whose complex, resource-intensive methods remain proprietary and opaque. This renders these efforts exclusive, leaving physicians dependent on them with limited autonomy^22^. In response, several open-source- friendly initiatives have developed resource-conscious approaches for domain adaptation, such as parameter-efficient tuning and retrieval-augmented generation (RAG). These approaches may provide alternative pathways without excessive computational and economic demands^23^. However, they still present technical challenges.

Here, we leverage a user-centered system to specialize LLM in EM through grounding methods aimed at enhancing the models’ contextual integration of queries and the verifiability of their responses. Using this system, we deployed closed- and open-source models to investigate their predictive diagnostic performance and sourcing capabilities in generating early diagnoses from limited, initial clinical data.

These predictions were compared to reference diagnoses established through expert consensus based on comprehensive, conclusive clinical data from a siloed, real-world dataset of a tertiary-care ED.

## Methods

### Study Design

This retrospective study was conducted at a tertiary ED in Belgium. We retrieved all admissions during the 24-hour peak influx period on a single day in 2023. Ethical approval was obtained from the institutional review board (ID 187/2023; Supplementary File S1), and all procedures adhered to applicable regulatory and ethical guidelines, including the Declaration of Helsinki and the EU’s General Data Protection Regulation 2016/679. All data were stored in a secure research database inaccessible to external entities, preventing data leakage and contamination into current or future AI training datasets.

### AI System

Neura^24^ deploys LLM with custom configurations and prompt engineering, harnessing curated corpora through advanced RAG^23^. For this study, we deployed two embedding models—text-embedding-ada-002 (OpenAI, San Francisco, US) and MXBAI (Mixedbread, Berlin, Germany)—with three foundational models—GPT-4 (OpenAI, San Francisco, US), Qwen2 (Alibaba, Hangzhou, China), and Llama3 (Meta, Menlo Park, US). A prototype corpus for clinical EM was curated, consisting of four textbooks: three on EM^25–27^ and one on critical care^28^.

### Evaluation Paradigm

Data were extracted from electronic health records to construct a comprehensive dataset, the ‘full clinical information,’ encompassing all clinical information recorded at the conclusion of each patient’s management in the ED. This dataset included patient demographics, vital signs, anamnesis (i.e., presenting complaints, past medical and surgical history, allergies, and current treatments), physical examination findings, as well as workup results and management decisions with procedural details from complementary examinations (i.e., examination type, location, findings, specialist involvement, and timestamps) of each case. In parallel, the conclusive ‘real-world diagnosis’ of the attending EP in clinical practice was recorded. The complete list of extracted parameters and definitions for each field is available in a data extraction sheet (Supplemental Table S1).

Three experienced EPs conducted an adjudication process to establish a ‘reference diagnosis’ for each case. First, two EPs independently reviewed the ‘full clinical information’ comprising the ‘real-world diagnosis.’ If they agreed, their consensus defined a consensual ‘reference diagnosis’; if they disagreed, the third EP resolved the case, determining a non-consensual ‘reference diagnosis.’

‘Early clinical information’ was defined as a subset of the ‘full clinical information,’ comprising only initial presentation parameters available at the onset of patient evaluation in the ED—i.e., anamnesis and physical examination findings. Based on this limited information, the AI was prompted to generate the top five most likely diagnoses, ranked by decreasing probability.

To evaluate AI performance, three independent investigators, distinct from the adjudicators and the attending EP, assessed whether any of the five AI-generated predictive diagnoses aligned with the reference diagnosis. For cases with correct alignment, they determined the rank of the matching diagnosis. Independently, the investigators classified each case as medical or surgical based on the ‘reference diagnosis.’ Additionally, each ‘reference diagnosis’ was categorized as either ’specific’ (i.e., ICD-10-specific) or ’unspecific’ (i.e., non-specific according to ICD-10 or outside the ICD-10 classification)^29^. Agreement between the two investigators confirmed the results for alignment, ranking, and classification (Figure 2).

**Figure 1.**
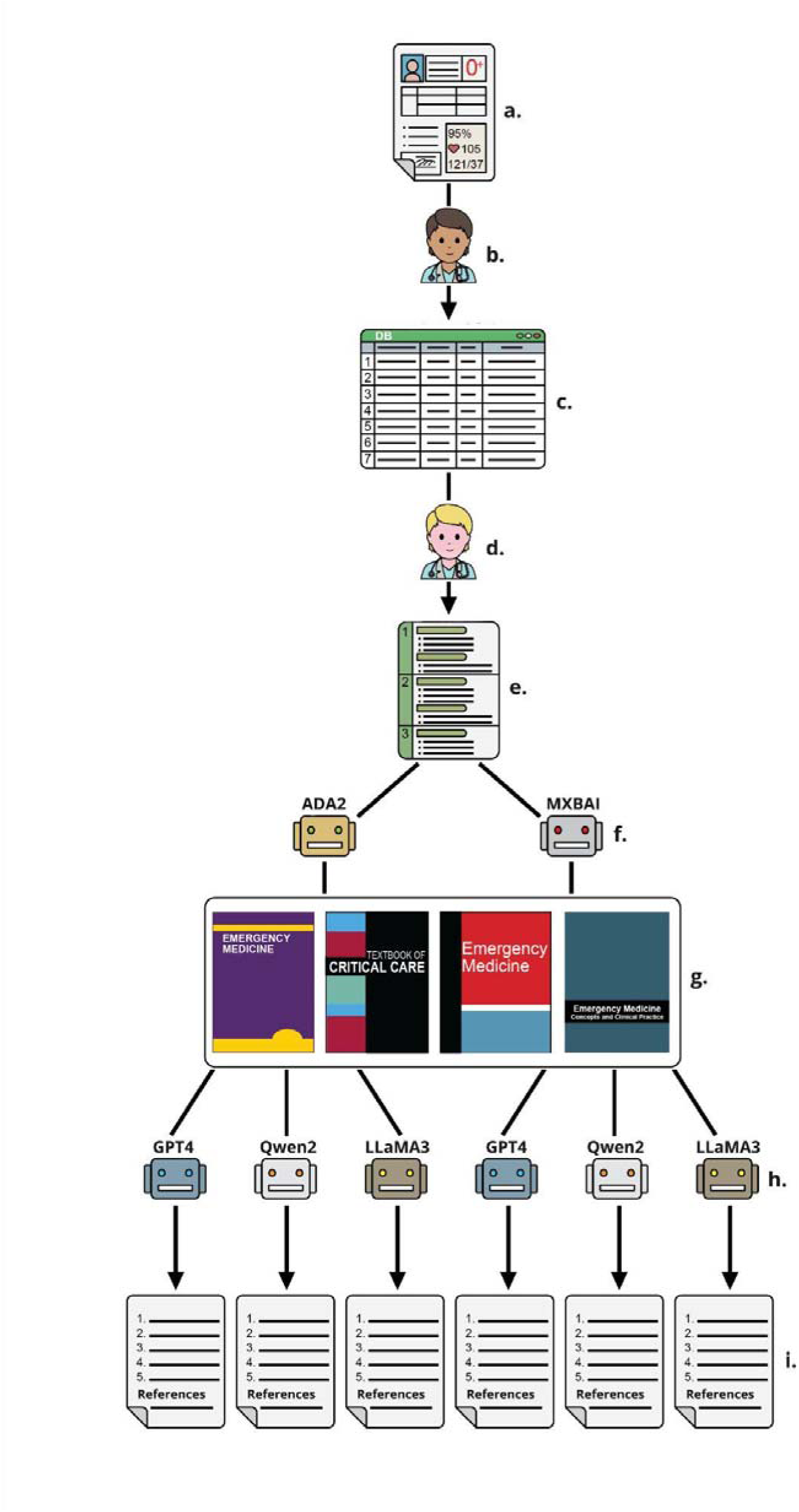
Data processing pipeline: **a.** Electronic health record (EHR) data; **b.** an independent investigator extracted clinical data from EHR; **c.** anonymized clinical data was stored in a structured database; **d.** an independent investigator prompted the models, **e.** using clinical vignettes; **f.** vignettes embedding using either the proprietary ADA2 model or the open-source MXBAI model**; g.** four EM textbooks constituted the curated corpus used for RAG; **h.** RAG output was processed by either GPT-4, Llama3, or Qwen2, were prompted the generation of the top five likely diagnoses, with each diagnosis aiming to be sourced from the relevant RAG knowledge base; **i.** top five diagnoses and references were generated for independent assessment.

**Figure 2.**
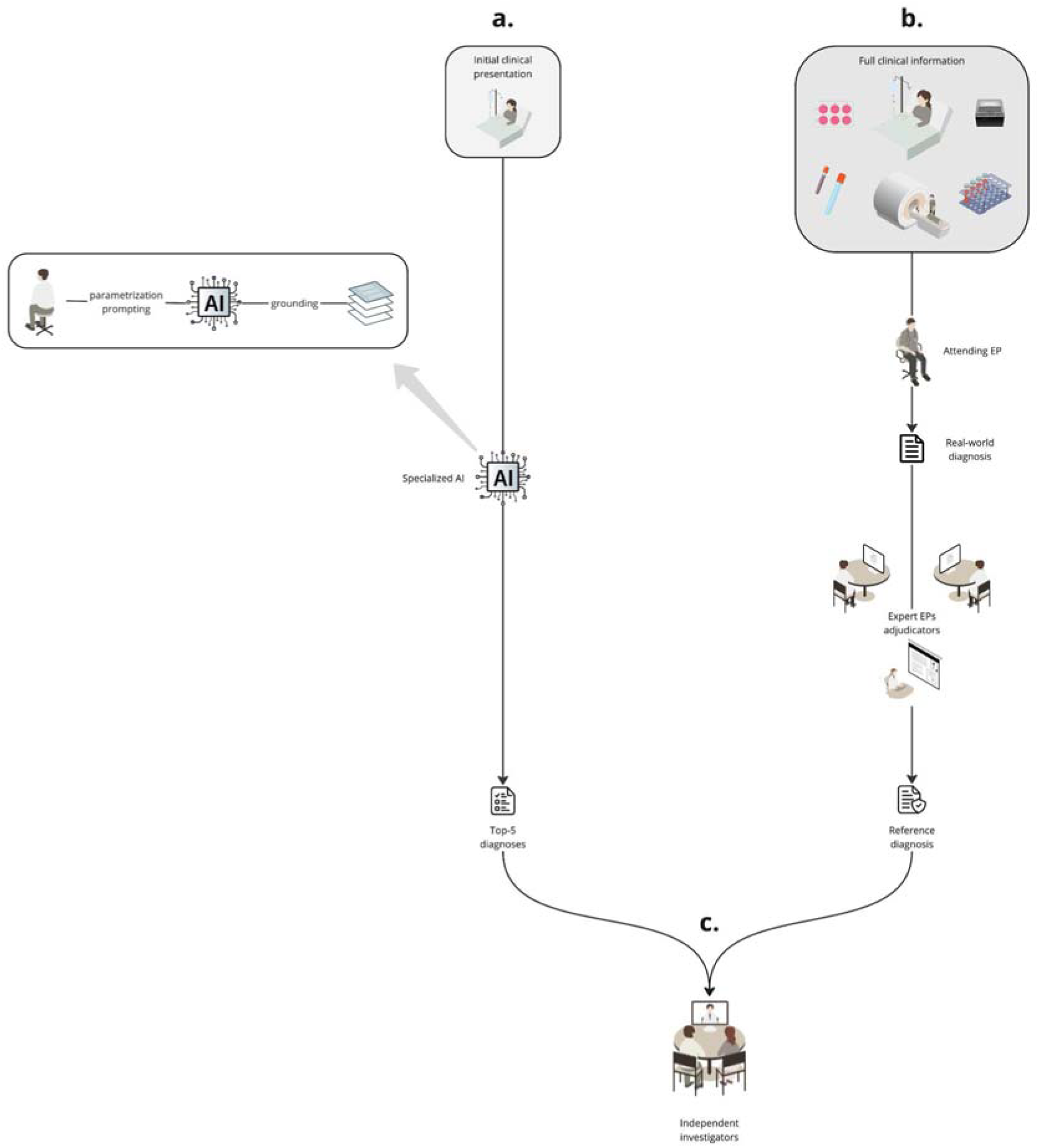
Diagnostic evaluation framework comparing AI-driven and clinical diagnostic pathways: **a.** AI pathway: Initial clinical presentation data is input into a specialized AI system that combines grounding techniques (RAG using emergency medicine textbooks), parametrization, and prompting to generate top-5 most likely diagnostic predictions. **b.** Clinical pathway: Full clinical information, including laboratory tests, imaging studies, and other diagnostic workup, is assessed by the attending EP to establish a real-world diagnosis, which is then independently reviewed by expert EP adjudicators to determine the reference diagnosis. **c.** Evaluation process: Independent investigators analyze the alignment between AI-generated diagnoses and the reference diagnosis.

### Statistical analysis

Diagnostic inclusion was defined as a binary outcome indicating whether the reference diagnosis appeared within the top five AI-generated predictions, and the inclusion rate was calculated as the proportion of cases with diagnostic inclusion out of the total number of cases. Diagnostic rank was measured on an ordinal scale (1- 5), representing the rank at which the reference diagnosis was provided—based on this position, we assigned 5 points for first place, decreasing by 1 point per rank down to 0 if absent and summed all points obtained per model combination across all cases to constitute a total ranking score. Diagnostic sourcing was assessed as a binary outcome, indicating whether verifiable citations substantiated the diagnosis.

Fisher’s exact test was used for categorical variables for pairwise comparisons, and the Mann-Whitney U test was employed to compare ordinal between two groups. To assess overall differences in ordinal across multiple groups, we used the Kruskal-Wallis test. The Chi-square test of independence was applied to evaluate associations between categorical variables. McNemar’s test was utilized for within-model comparisons of paired categorical data.

We implemented multiple regression models tailored to each outcome’s characteristics.

For binary outcomes (diagnostic inclusion and sourcing), we employed logistic regression models:

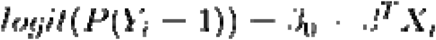

where Yi represents the binary outcome, Xi is the vector of predictors, β0 is the intercept, and βT is the transpose of the vector of regression coefficients. Models were fitted using maximum likelihood estimation with LBFGS optimization (maximum 1,000 iterations), with Newton’s method as a fallback optimization strategy. Model performance was assessed through multiple metrics, including classification accuracy, precision (positive predictive value), recall (sensitivity), F1-score, and ROC curves with AUC calculations.

For analyzing ranking performance, we developed an ordinal score transformation:

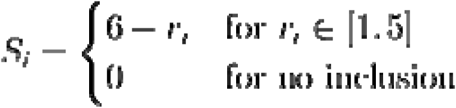

where ri represents the original rank position and Si is the transformed score. The relationship between predictors and ranking performance was modeled using ordinary least squares regression:

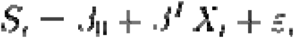

where εi represents the error term, assumed to be normally distributed with constant variance.

All models underwent diagnostic testing. We calculated McFadden’s pseudo-R² for binary outcomes and conducted likelihood ratio tests. For the rank analysis, we examined model fit through R² and adjusted R² coefficients, F-statistics with associated p-values, Durbin-Watson statistic for autocorrelation, Jarque-Bera test for normality of residuals, White’s test for heteroscedasticity and Rainbow test for linearity of relationships.

Multicollinearity was assessed using variance inflation factors (VIF).

To quantify the relative contribution of each predictor to model performance, we calculated standardized importance measures:

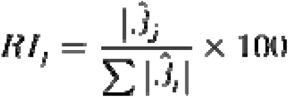

where βj represents the estimated coefficient for predictor j, and RIj indicates its relative importance percentage.

Residual diagnostics were visualized using residual histograms and residuals vs. fitted values, normal Q-Q, and scale-location plots.

For binary outcomes, ROC curves were generated to visualize the trade-off between sensitivity and specificity across different classification thresholds.

Python 3.10 was employed using pandas (v1.5.3) for data manipulation, scipy (v1.10.1) for statistical testing, matplotlib (v3.7.1), seaborn (v0.12.2) for data visualization, and statsmodels (v0.14.0) for statistical modeling. Statistical significance was set at α = 0.05 for all analyses.

## Results

In 2023, the peak ED influx occurred on a summer day. We retrieved 79 cases, of which 53 were classified as specific, 61 as consensual and 35 as medical. Table 1 summarizes the characteristics of the cohort. Figure 3 displays the variety of diagnostic codes according to ICD-10.

**Figure 3.**
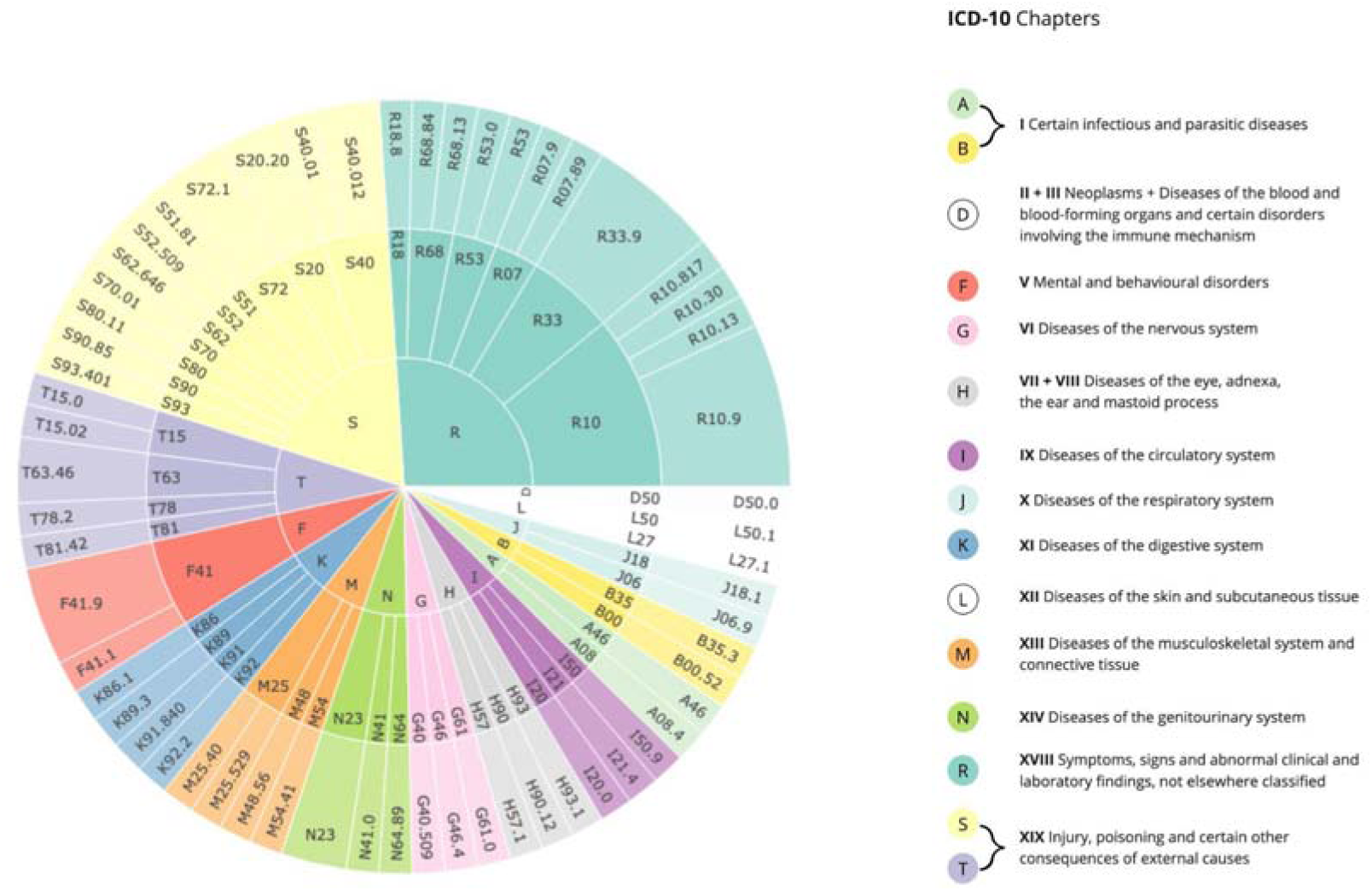
Distribution of International Classification of Diseases, 10th Revision (ICD-10) diagnostic codes. Sunburst diagram shows the hierarchical organization of diagnoses by ICD-10 chapters (letters A-T) and specific diagnostic codes. Color-coded sections represent major ICD-10 chapters.

**Table 1.** Cases characteristics.

| Characteristics | n (%) |
| --- | --- |
| **Patients demographics (n=79)** |  |
| Sex |  |
| Male | 45 (57.0) |
| Female | 34 (43.0) |
| Age, median (IQR), years | 57 (43-73) |
| **Case classification (n=79)** |  |
| Specific diagnoses | 53 (67.1) |
| Unspecific diagnoses | 26 (32.9) |
| **Medical discipline (n=79)** |  |
| Medical | 35 (44.3) |
| Surgical | 39 (49.4) |
| Psychiatric | 5 (6.3) |
| **Adjudication (n=79)** |  |
| Consensual cases | 61 (77.2) |
| Non-consensual cases | 18 (22.8) |
| **Diagnostic categorization (n=79)** |  |
| ICD-10 coded diagnoses | 73 (92.4) |
| Non-ICD-10 coded diagnoses | 6 (7.6) |
| **Specialty distribution (n=79)** |  |
| Orthopedics | 29 (36.7) |
| Gastroenterology | 12 (15.2) |
| Internal Medicine | 7 (8.9) |
| Urology | 7 (8.9) |
| Neurology | 6 (7.6) |
| Psychiatry | 5 (6.3) |
| Ophthalmology | 4 (5.1) |
| Dermatology | 4 (5.1) |
| ENT | 3 (3.8) |
| Cardiology | 2 (2.5) |
| Others* | 5 (6.3) |

\*Others include Stomatology (2), Pneumology (2), Vascular Surgery (1), Plastic Surgery (1), and Pediatrics (1).

For diagnostic inclusion, model combinations demonstrated rates ranging from 62.03% to 72.15%, with no statistically significant differences in pairwise comparisons (all p>0.05, Fisher’s exact tests).

In subgroup analyses, we found that (i) specific cases showed significantly higher inclusion versus unspecific cases (85.53% vs. 31.41%, OR=13.00, p<0.001), a pattern consistent across all model combinations (all p<0.001); (ii) surgical cases had significantly higher inclusion than medical cases (79.49% vs. 56.25%, OR=3.01, p=5.74e-08), with this difference reaching statistical significance at the individual model level for ADA/GPT-4 and MBXAI/LLaMA3 combinations (OR=3.69-4.13, p=0.008-0.010); (iii) consensual cases achieved significantly higher inclusion versus non-consensual cases (71.58% vs. 54.63%, OR=2.092, p=0.001), though this difference was not statistically significant at the individual model level (all p>0.05) (Figure 4).

**Figure 4.**
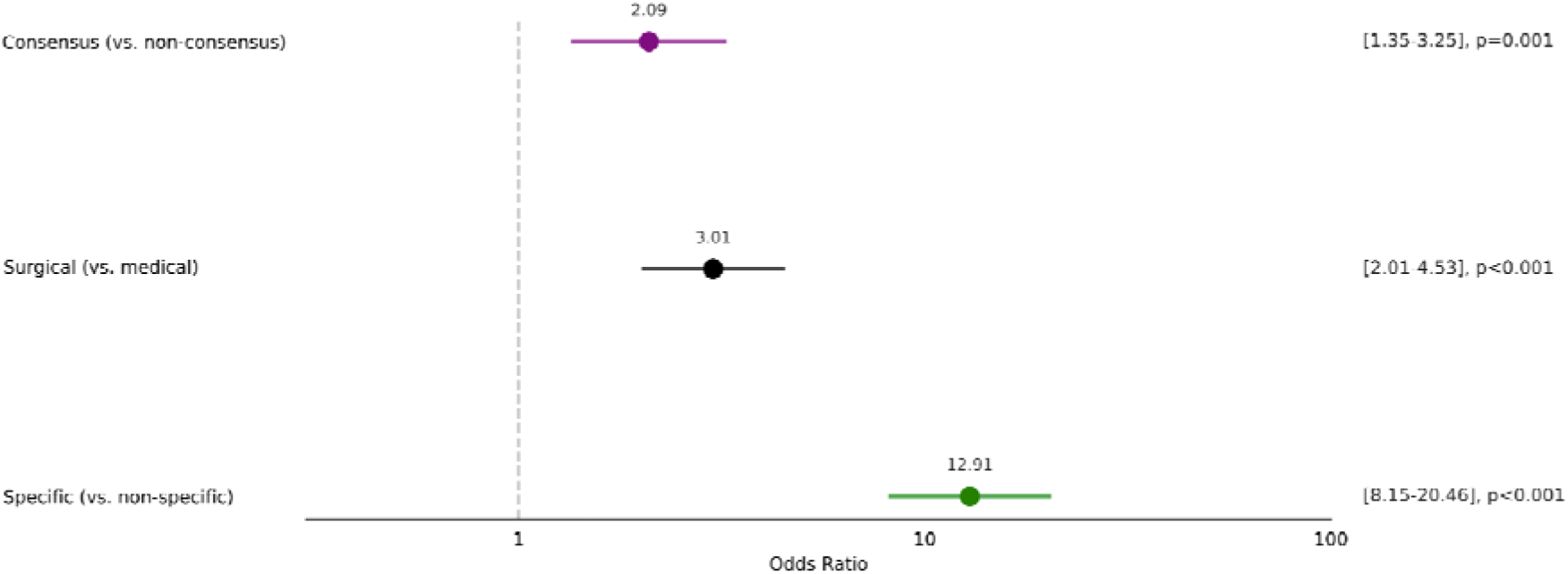
Forest plot of odds ratios for diagnostic inclusion stratified by case characteristics. Points represent odds ratios with 95% confidence intervals (horizontal lines) on a logarithmic scale. Comparisons shown are specific versus non-specific diagnoses (green), surgical versus medical cases (black), and consensus versus non-consensus cases (purple). Values to the right of the vertical dashed line (OR > 1) indicate higher odds of diagnostic inclusion.

Notably, pairwise comparisons between model combinations within each subgroup (i.e., specific/unspecific, medical/surgical, consensual/non-consensual) revealed no significant differences in diagnostic inclusion (Fisher’s exact tests).

In the subgroup analysis of cases with poor inclusion performance, we found that (i) null-inclusion cases (n=16) showed a significant predominance of unspecific diagnoses (87.5%, OR=33.33, p<0.0001) and medical presentations (75%, OR=3.75, p=0.0482), while consensus rates remained comparable; (ii) mixed- inclusion cases (n=25) showed no significant differences in characteristics (all p>0.05, Fisher’s exact tests) (Figure 5).

**Figure 5.**
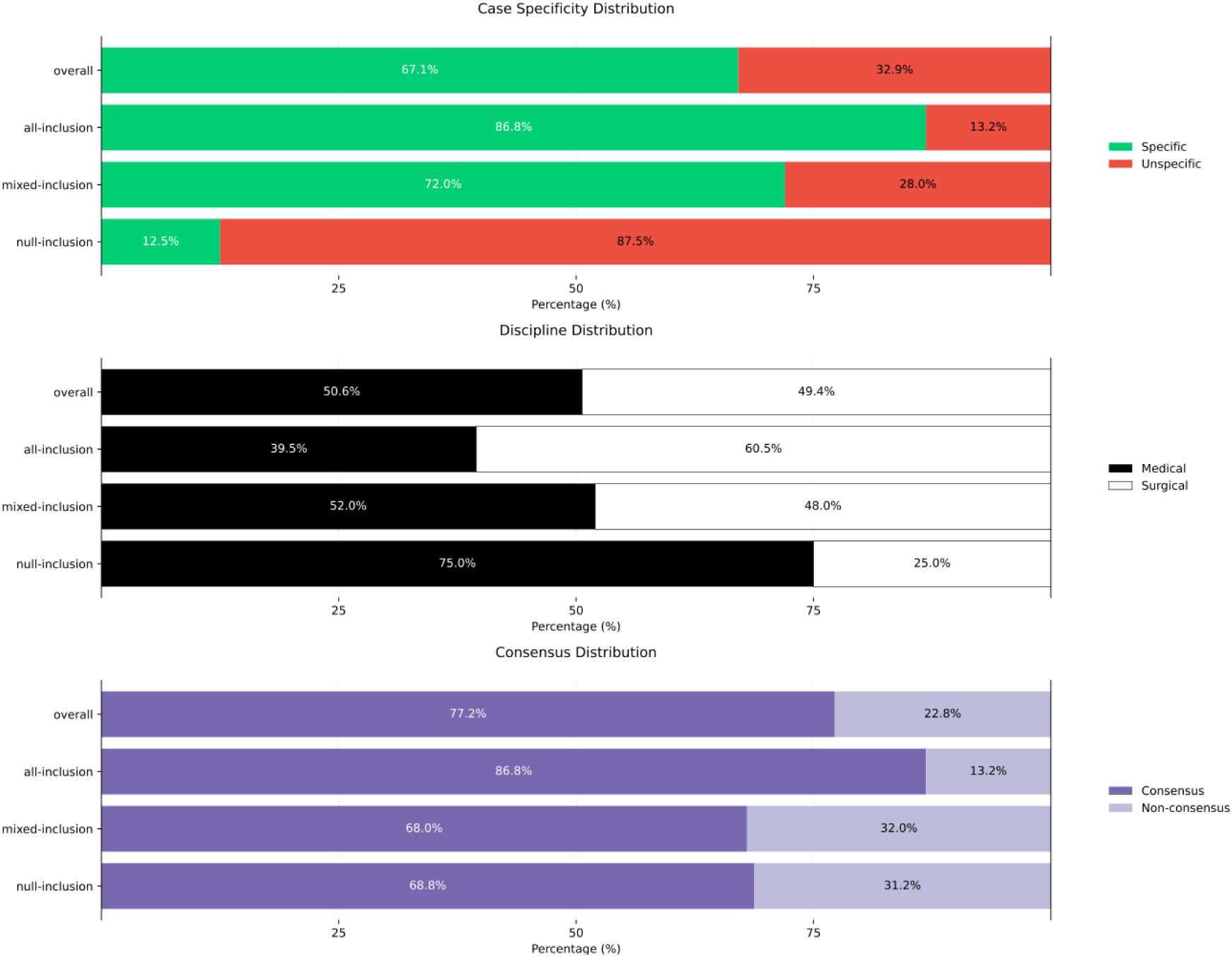
Distribution of case characteristics stratified by diagnostic inclusion performance. Horizontal stacked bar plots show the percentage distribution of (A) case specificity (specific vs. unspecific diagnoses, green/red), (B) medical discipline (medical vs. surgical cases, black/white), and (C) diagnostic consensus (consensus vs. non-consensus cases, dark/light purple). Cases are categorized by inclusion performance: null-inclusion (no models included the reference diagnosis), mixed-inclusion (some models included the reference diagnosis), all-inclusion (all models included the reference diagnosis), and overall distribution across all cases.

Ranking analysis showed total scores ranging from 214 to 245 points and mean ranks between 1.01 and 1.39, with no significant differences in distribution across model combinations (Kruskal-Wallis test) or pairwise comparisons (Mann-Whitney U tests).

Regarding diagnostic sourcing, LLaMA3-based and QWEN2-based combinations demonstrated significantly higher rates compared to GPT-4-based combinations (OR: 33.92 to ∞ for MBXAI/QWEN2 achieving perfect sourcing rate; p<1.4e-12), with no other significant differences between model combinations (Fisher’s exact tests) (Figure 6).

**Figure 6.**
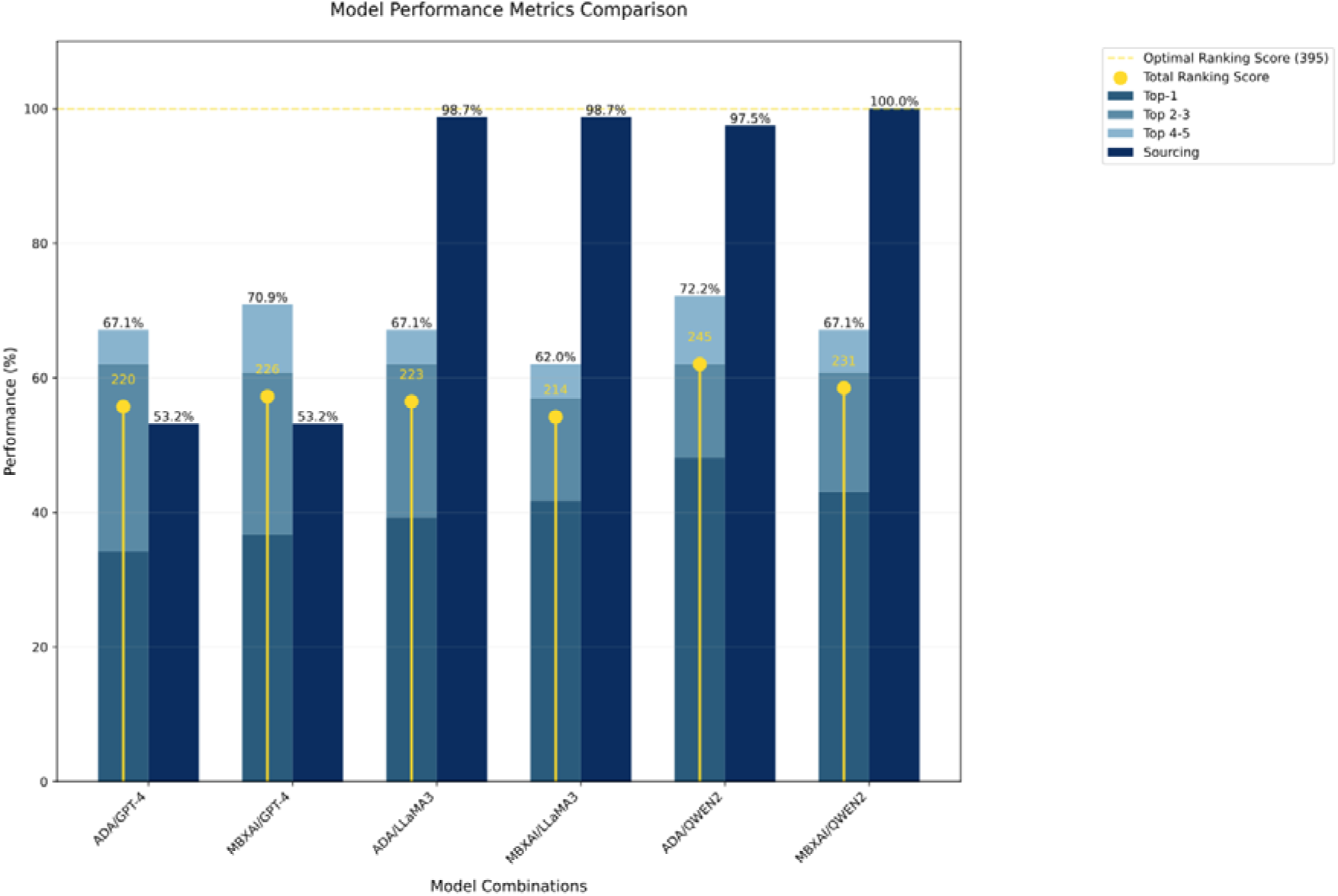
Performance metrics comparison across six model combinations. Stacked bar chart displays diagnostic performance metrics for each embedding/foundation model combination. Darkest bars show Top-1 inclusion rates (percentage of cases where the reference diagnosis was the model’s top prediction), medium bars show Top-3 inclusion rates, and lightest bars show Top-5 inclusion rates (percentage of cases where the reference diagnosis appeared among the model’s top five predictions). Dark navy bars represent sourcing rates (percentage of cases with verifiable citations). Yellow dots indicate total ranking scores, with the optimal possible score (395) shown by the horizontal dashed line.

For diagnostic inclusion, the logistic regression model demonstrated significant fit (χ² = 174.46, p = 5.088e-35) with a McFadden’s pseudo-R² of 0.2926, log-likelihood of -210.89 (null: -298.12), degrees of freedom of 467 (residual) and 6 (model), and constant term (β[= 0.9246, SE = 0.334, p = 0.006), revealing significant effects for consensus status (β = 0.7192, SE = 0.270, 95% CI [0.191, 1.248], p = 0.008), specific diagnosis (β = 2.6642, SE = 0.253, 95% CI [2.168, 3.161], p < 0.001), and surgical discipline (β = 1.2303, SE = 0.256, 95% CI [0.728, 1.732], p < 0.001), while model comparisons showed no significant impact: MBXAI vs ADA (β = -0.1488, SE = 0.244, 95% CI [-0.627, 0.330], p = 0.542), LLaMA3 vs GPT-4 (β = -0.3084, SE = 0.298, 95% CI [-0.892, 0.275], p = 0.300), and QWEN2 vs GPT-4 (β = 0.0452, SE = 0.302, 95% CI [-0.546, 0.637], p = 0.881); relative importance analysis identified specific diagnosis (52.07%) and surgical discipline (24.05%) as the strongest predictors. The class distribution showed 67.72% positive cases with strong model performance metrics (accuracy=0.812, precision=0.835, recall=0.900, F1- score=0.867). The information criteria were AIC=435.784 and BIC=464.913.

For diagnostic ranking, the linear regression model (F = 29.83, p = 2.70e-30) achieved R² = 0.277 (adjusted R² = 0.268), with log-likelihood of -967.38, degrees of freedom of 467 (residual) and 6 (model)], with constant term (β[= 2.8361, SE = 0.256, p < 0.001) and significant coefficients for consensus status (β = 0.6289, SE = 0.207, 95% CI [0.222, 1.036], p = 0.003), specific diagnosis (β = 2.2012, SE = 0.185, 95% CI [1.838, 2.564], p < 0.001), and surgical discipline (β = 0.5296, SE = 0.174, 95% CI [0.187, 0.872], p = 0.003), while model combinations variables remained non-significant (MBXAI vs ADA: β = -0.0717, SE = 0.172, p = 0.678; LLaMA3 vs GPT-4: β = -0.0570, SE = 0.211, p = 0.787; QWEN2 vs GPT-4: β = 0.1899, SE = 0.211, p = 0.369); the model satisfied diagnostic criteria (Durbin-Watson = 2.047, condition number = 6.16) with a slight departure from normality (skewness = -0.196, kurtosis = 2.482, Jarque-Bera = 8.353, p = 0.0154). The rank score distribution showed bimodal characteristics with peaks at score 0 (n=153) and score 5 (n=192). Residual analysis for the rank model demonstrated zero-centered residuals (mean=0.000, SD=1.863) with slight negative skewness (-0.196) and platykurtic distribution (kurtosis=-0.518); linearity assumption was supported by the Rainbow test (p=0.777), and independence was confirmed by the Durbin-Watson statistic (2.047). Multicollinearity assessment through VIF showed consistent values across all models, with minimal collinearity among predictors (consensus: 1.014, unspecific: 1.012, embedding model being MBXAI: 1.600, foundational model being LLaMA3 or QWEN2: 1.400, surgical discipline: 1.616). Influence diagnostics revealed well-controlled leverage (maximum=0.023, mean=0.015) and Cook’s distance values (maximum=0.015, mean=0.002), indicating no influential outliers. White’s test for heteroscedasticity showed borderline results (LM test: p = 0.05021; F-test: p = 0.04463), suggesting no severe violation of the homoscedasticity assumption and information criteria were: AIC=1949, BIC=1978.

Regarding diagnostic sourcing, the logistic regression achieved McFadden’s pseudo- R² of 0.3842 (χ² = 162.84, p = 1.478e-32), log-likelihood of -130.53 (null: -211.95), and degrees of freedom of 467 (residual) and 6 (model), with constant term (β[= 0.1074, SE = 0.401, p = 0.789), revealing significant positive effects for LLaMA3 vs. GPT-4 and QWEN2 vs. GPT-4 (both β = 4.2323, SE = 0.729, 95% CI [2.803, 5.662], p < 0.001), which dominated relative importance (48.11% each), while other comparisons showed no significant effects (consensus status: β = -0.1052, SE = 0.367, p = 0.775; specific diagnosis: β = -0.0708, SE = 0.326, p = 0.828; MBXAI vs ADA: β = 0.0924, SE = 0.304, p = 0.761; surgical discipline: β = 0.0635, SE = 0.308, p = 0.836). The class distribution revealed 83.54% positive cases with robust performance metrics (accuracy=0.835, precision=0.835, recall=1.000, F1- score=0.910). The information criteria were AIC=275.053 and BIC=304.181.

All three models demonstrated adequate fit metrics, with pseudo-R² values of 0.293 and 0.384 for binary outcomes (top-5 and sourcing, respectively) and R²=0.277 (adjusted R²=0.268) for the rank model.

## Discussion

In this real-world evaluation, we found that both open- and closed-source models, grounded with domain-specific knowledge, achieve comparable predictive performance with overall diagnostic inclusion rates ranging from 62% to 72%.

Pairwise comparisons indicated no significant differences in diagnostic inclusion or ranking performance among the various combinations of embedding and foundation models. Moreover, when models successfully included a diagnosis, they consistently ranked it among their top proposals with mean ranks between 1 and 1.4. Hence, the disproportionate representation of proprietary commercial systems in the biomedical literature reporting LLM-driven diagnosis likely stems from transient productization advantages rather than intrinsic technological superiority^12^. Along these lines, our study demonstrates that diagnostic performance here was primarily determined by case characteristics rather than model combinations, as evidenced by our regression analysis identifying specific diagnosis and surgical discipline as the strongest predictors with 52 and 24% of relative importance. Specifically, ICD-10-specific diagnoses, as well as surgical and consensual cases, demonstrated higher diagnostic inclusion, ranging from 71% to 85%—this pattern was consistently mirrored in ranking performance. Notably, our analysis of the 16 null-inclusion cases revealed a significant predominance of 87.5% unspecific diagnoses and 75% medical presentations. This convergent behavior suggests that current limitations in LLM-driven diagnosis reflect fundamental AI alignment challenges in medical reasoning rather than the technological limitations of specific models. Lastly, open-source foundational models showed significantly superior sourcing capabilities, underscoring distinct advantages in clinical decision support that build upon their inherent alignment with greater transparency and verifiability.

As AI performance patterns across different case types demonstrate how technology adapts to the complexities of medical diagnosis, addressing these challenges requires us to go beyond mere technical metrics. We must critically assess diagnostic reasoning, recognize its inherent uncertainty and nonspecificity, and create clinically relevant solutions. Additionally, we should explore how various clinical scenarios might support or confront these systems. Yet, medicine lacks a unified, unequivocal clinical code—a longstanding challenge predating the advent of biomedical AI^30–33^. Against this complex backdrop, evaluating AI diagnostic capabilities requires precise definitional frameworks and robust comparative standards, which are currently lacking^34^. For instance, the poor model performance on unspecific diagnoses suggests that ambiguity in clinical definition and communication should be acknowledged, studied, and mitigated rather than simply viewed as a failure of either artificial or human intelligence. Here, we adopted an AI-generated top-five most likely diagnoses paradigm as a dual proxy for (i) the differential diagnosis process of EPs and (ii) a clinically relevant contribution to this endeavor. Of course, this simplistic paradigm cannot fully capture the gestalt of EPs’ differential diagnosis. Moreover, in emergency medicine, the rapid identification of relevant diagnoses based on limited information occurs in a continuously dynamic assessment, where identifying or ruling out time-sensitive and potentially severe conditions often take precedence over diagnostic probability^5,35,36^. We also employed a structured adjudication process involving three experienced EPs who reviewed full clinical data to establish a final reference diagnosis. As this approach allows validation against ground truth, it differs from identifying relevant diagnoses at any given moment—a process that may encompass a broad range of possibilities, confine to subjectivity, and ultimately diverge from practical clinical endpoints^37,38^.

In this study, multi-level grounding approaches could achieve verifiable predictions without prohibitive computational resources by combining RAG with parameterization and prompting. These benefits manifested particularly in open-source architectures, which demonstrated marked superiority in sourcing capabilities, LLaMA3-based and Qwen2-based pipelines displaying substantially higher rates of verifiable references than GPT-4-based ones, with one open-source combination, MBXAI/Qwen2, achieving a perfect sourcing rate. Interestingly, embedding models showed no significant influence on sourcing, suggesting that proprietary API deployments of foundational models involve opaque back-end processing that may impede custom grounding. This lack of transparency extends beyond technical considerations but involves ethical and regulatory issues on model interpretability and accountability in clinical settings. In emergency medicine, where decisions must be both rapid and defensible, verification of AI recommendations against a curated knowledge base enables controlled and effective clinical oversight. This transparency reinforces physician autonomy by providing direct control over AI assistance while establishing clear lines of medical responsibility and liability^39–41^. However, providing sources does not ensure their verification in practice—a time-consuming and cognitively demanding task—potentially making sourcing features counterproductive by fostering unwarranted trust. In parallel, the mere use of AI does not inherently enhance physician diagnostic reasoning^42,43^. This reality demands thoughtful integration of AI through intuitive interfaces that guide optimal human-computer interactions (Figure 7) alongside semi-automated LLM-driven reference verification^43,44^. Such verifiability, while essential for maintaining physician agency, must extend beyond technical features to promote appropriate trust and build scientific and technological literacy—crucial elements in an increasingly AI-augmented clinical environment.

**Figure 7.**
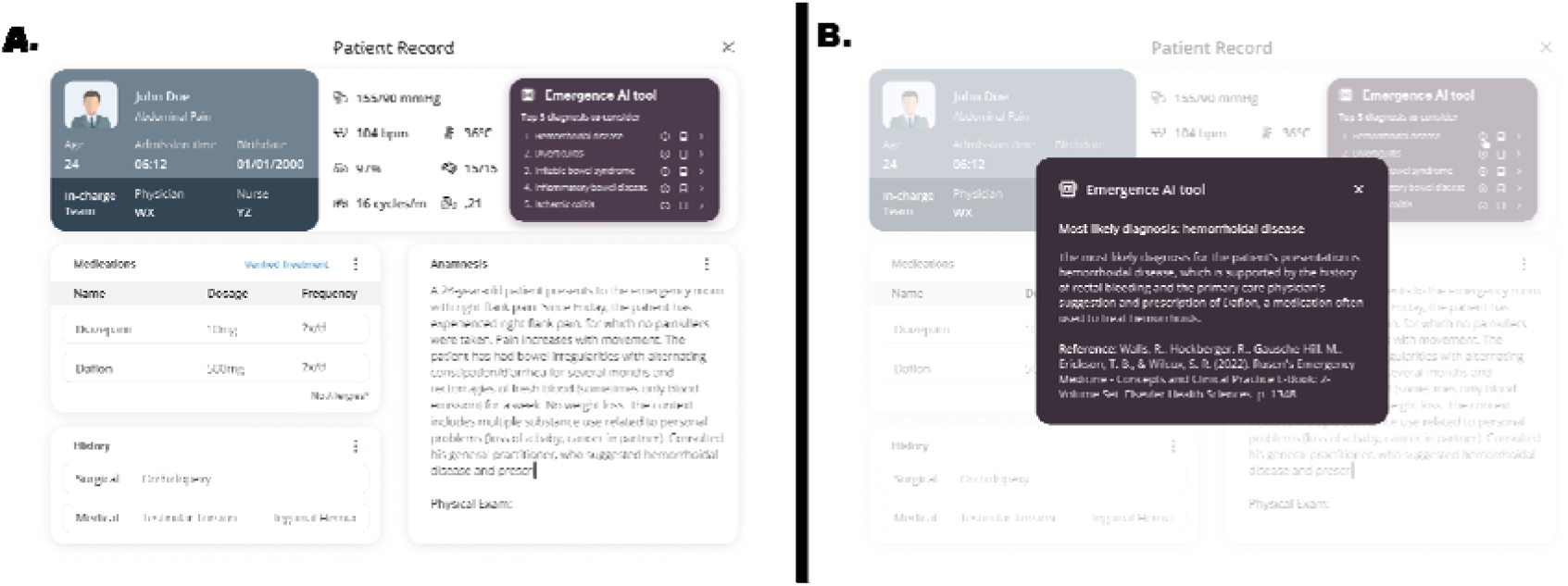
User interface example for AI-assisted diagnostic support. **A.** Initial view showing patient electronic health record with vital signs and summary interface for the AI diagnostic tool displaying top 5 diagnoses to consider. **B.** Expanded view showing detailed explanation of the most likely diagnosis with supporting clinical reasoning and reference citation from literature.

## Limitations

While we utilized real-world, siloed ED data from a tertiary center amid a peak influx of patients and applied an independent adjudication framework to strengthen ecological validity and methodological robustness, several limitations persist. First, the monocentric and retrospective design during peak 24-hour limits broader generalizability. Second, although using clinical vignettes mimics a natural form of medical communication, this step inevitably introduces the risk of language biases. Third, we deployed RAG on a prototype curated corpus consisting of only four textbooks. Lastly, Llama 3 was originally restricted to an 8,192-token context window therefore requiring positional embedding interpolation^45^—which may have impacted performance compared to Qwen2 and GPT-4, both of which natively support 128,000 tokens. Since then, Meta’s Llama 3.1 and 3.2 with extended context windows, along with newer OpenAI’s series of reflective models with integrated chain-of-thought and Alibaba’s Qwen2.5, have been released—each potentially altering relative performance if re-evaluated under similar conditions.

### Perspectives

Future research will benefit from prospective, multicenter, and multilingual trials that evaluate LLM-driven diagnostic support in diverse practice settings across larger and more heterogeneous patient populations. Integrating iterative data updates—e.g., findings from laboratory and imaging examinations, specialist consultations, and clinical evolution—into real-time LLM-driven diagnosis would better emulate the dynamic nature of emergency medicine. We should examine how LLM-based tools streamline ED triage by supporting physician decision-making. Critical outcomes include the prioritization and acceleration of critical care delivery for time-sensitive and high-risk cases while facilitating the primary care of benign conditions— specifically measuring how these tools enhance the quality of care provided but also reduce, rather than add to, physician cognitive load and administrative burden^46–48^. Furthermore, expanding and specifying the AI knowledge base—such as by incorporating hospital guidelines or regional epidemiology—could enhance the relevance of such diagnostic support. In this regard, open-source approaches hold particular promise for on-premises deployments. They may mitigate privacy risks and align with strict data-sharing regulations. Building upon these advantages, purpose-built LLM-based agentic infrastructure could contribute to patient management workflows—from triage and clinical assessment to treatment planning and discharge—with explicit reasoning traces and self-correction capabilities, enabling transparent processes that adapt to evolving clinical information while maintaining accountability across healthcare providers’ interactions^49,50^. These solutions could offer a viable path for institutions seeking technological independence with granular control over data governance and model interpretability.

## Conclusion

In this real-world ED setting, open- and closed-source LLM pipelines demonstrated promising and comparable performance in generating top-5 predictive diagnoses. Case characteristics emerged as the dominant influence of diagnostic performance—elucidating AI alignment challenges extending beyond technological optimization to fundamental aspects of clinical reasoning patterns. Notably, open-source pipelines demonstrated significantly superior sourcing capabilities, an important consideration for interpretable AI solutions. As emergency departments contend with heightened patient volumes and staffing shortages, such specialized AI-driven tools may become a valuable asset for supporting timely, well-informed clinical decision-making. Continued research exploring larger-scale, multi-centric efforts, including real-time applications and human-computer interactions, as well as benchmarking against clinical outcomes, workflow efficiency, and reference verification, will be key to delineating the full potential of grounded LLM-driven diagnostic assistance in emergency medicine.

## Data Availability

We have elected not to make our original patient-level dataset publicly available. As this dataset originates from real-world emergency department cases, it contains sensitive clinical information despite de-identification. We are particularly concerned that sharing these data may lead to unintended contamination and dissemination among external models used for training purposes. Such dissemination could compromise the integrity of the dataset and preclude its future use in further research. Consequently, we have decided to restrict access to the data to preserve its unique value and to ensure that its subsequent application in research remains controlled and ethical.

## Bibliography

1. Varner C. Emergency departments are in crisis now and for the foreseeable future. CMAJ 2023;195(24):E851–E852. DOI: 10.1503/cmaj.230719.

2. Physicians ACoE. Letter to President Biden. 2022.

3. Kellermann AL. Crisis in the emergency department. N Engl J Med 2006;355(13):1300–3. DOI: 10.1056/NEJMp068194.

4. Clancy CM. Emergency departments in crisis: opportunities for research. Health Serv Res 2007;42(1 Pt 1):xiii-xx. DOI: 10.1111/j.1475-6773.2006.00692.x.

5. Kovacs G, Croskerry P. Clinical decision making: an emergency medicine perspective. Acad Emerg Med 1999;6(9):947–52. DOI: 10.1111/j.1553-2712.1999.tb01246.x.

6. Chenais G, Lagarde E, Gil-Jardine C. Artificial Intelligence in Emergency Medicine: Viewpoint of Current Applications and Foreseeable Opportunities and Challenges. J Med Internet Res 2023;25:e40031. DOI: 10.2196/40031.

7. Brown TB, Mann B, Ryder N, et al. Language Models are Few-Shot Learners. (https://ui.adsabs.harvard.edu/abs/2020arXiv200514165B).

8. Liu N, Chen L, Tian X, Zou W, Chen K, Cui M. From LLM to Conversational Agent: A Memory Enhanced Architecture with Fine-Tuning of Large Language Models. (https://ui.adsabs.harvard.edu/abs/2024arXiv240102777L).

9. Mintz Y, Brodie R. Introduction to artificial intelligence in medicine. Minim Invasive Ther Allied Technol 2019;28(2):73–81. DOI: 10.1080/13645706.2019.1575882.

10. Mesko B, Topol EJ. The imperative for regulatory oversight of large language models (or generative AI) in healthcare. NPJ Digit Med 2023;6(1):120. DOI: 10.1038/s41746-023-00873-0.

11. Bommasani R, Hudson DA, Adeli E, et al. On the Opportunities and Risks of Foundation Models. (https://ui.adsabs.harvard.edu/abs/2021arXiv210807258B).

12. Li J, Dada A, Puladi B, Kleesiek J, Egger J. ChatGPT in healthcare: A taxonomy and systematic review. Comput Methods Programs Biomed 2024;245:108013. DOI: 10.1016/j.cmpb.2024.108013.

13. Kung TH, Cheatham M, Medenilla A, et al. Performance of ChatGPT on USMLE: Potential for AI-assisted medical education using large language models. PLOS Digit Health 2023;2(2):e0000198. DOI: 10.1371/journal.pdig.0000198.

14. Thirunavukarasu AJ, Ting DSJ, Elangovan K, Gutierrez L, Tan TF, Ting DSW. Large language models in medicine. Nature medicine 2023;29(8):1930–1940. DOI: 10.1038/s41591-023-02448-8.

15. Lee P, Bubeck S, Petro J. Benefits, Limits, and Risks of GPT-4 as an AI Chatbot for Medicine. N Engl J Med 2023;388(13):1233–1239. DOI: 10.1056/NEJMsr2214184.

16. Commission E. Proposal for a REGULATION OF THE EUROPEAN PARLIAMENT AND OF THE COUNCIL LAYING DOWN HARMONISED RULES ON ARTIFICIAL INTELLIGENCE (ARTIFICIAL INTELLIGENCE ACT) AND AMENDING CERTAIN UNION LEGISLATIVE ACTS. In: European Parliament CotEU, ed. Brussels2021.

17. Union E. Regulation (EU) 2016/679 of the European Parliament and of the Council of 27 April 2016 on the protection of natural persons with regard to the processing of personal data and on the free movement of such data, and repealing Directive 95/46/EC (General Data Protection Regulation). In: European Parliament CotEU, ed. 02016R0679-20160504. online2016.

18. SERVICES DOHAH. Modifications to the HIPAA Privacy, Security, Enforcement, and Breach Notification Rules Under the Health Information Technology for Economic and Clinical Health Act and the Genetic Information Nondiscrimination Act; Other Modifications to the HIPAA Rules In: Office for Civil Rights DoHaHS, ed. Online2013.

19. OpenAI, Achiam J, Adler S, et al. GPT-4 Technical Report. (https://ui.adsabs.harvard.edu/abs/2023arXiv230308774O).

20. Carlini N, Tramer F, Wallace E, et al. Extracting Training Data from Large Language Models2020.

21. Gupta M, Akiri C, Aryal K, Parker E, Praharaj L. From ChatGPT to ThreatGPT: Impact of Generative AI in Cybersecurity and Privacy. IEEE Access 2023;11:80218–80245. DOI: 10.1109/access.2023.3300381.

22. Lin P, Tao H, Li H, Huang S-Y. Protein–protein contact prediction by geometric triangle-aware protein language models. Nature Machine Intelligence 2023;5(11):1275–1284. DOI: 10.1038/s42256-023-00741-2.

23. Lewis P, Perez E, Piktus A, et al. Retrieval-Augmented Generation for Knowledge-Intensive NLP Tasks. (https://ui.adsabs.harvard.edu/abs/2020arXiv200511401L).

24. Barrit S, Torcida N, Mazeraud A, et al. Neura: a specialized large language model solution in neurology. medRxiv 2024:2024.02.11.24302658. DOI: 10.1101/2024.02.11.24302658.

25. Tintinalli JE, Stapczynski JS, Ma OJ, Yealy DM, Meckler GD, Cline DM. Tintinalli’s Emergency Medicine: A Comprehensive Study Guide, 8e. New York, NY: McGraw-Hill Education; 2016.

26. Walls RM, Hockberger RS, Gausche-Hill M, Erickson TB, Wilcox SR. Rosen’s Emergency Medicine: Concepts and Clinical Practice: Elsevier, 2022.

27. Wyatt JP, Taylor RG, de Wit K, Hotton EJ. Oxford Handbook of Emergency Medicine2020.

28. Vincent JL, Moore FA, Bellomo R, Marini JJ. Textbook of Critical Care E-Book: Elsevier, 2022.

29. World Health O. ICD-10 : international statistical classification of diseases and related health problems : tenth revision. 2nd ed. Geneva: World Health Organization; 2004.

30. Feinstein AR. The problems of the “problem-oriented medical record”. Ann Intern Med 1973;78(5):751–62. DOI: 10.7326/0003-4819-78-5-751.

31. Weed LL. Medical records that guide and teach. N Engl J Med 1968;278(11):593–600. DOI: 10.1056/NEJM196803142781105.

32. Cimino JJ. Desiderata for controlled medical vocabularies in the twenty-first century. Methods Inf Med 1998;37(4-5):394–403. (In eng) (https://www.ncbi.nlm.nih.gov/pubmed/9865037).

33. Rector AL, Solomon WD, Nowlan WA, Rush TW, Zanstra PE, Claassen WM. A Terminology Server for medical language and medical information systems. Methods Inf Med 1995;34(1-2):147–57. (In eng) (https://www.ncbi.nlm.nih.gov/pubmed/9082124).

34. Johnson R, Gottlieb U, Shaham G, et al. Unified Clinical Vocabulary Embeddings for Advancing Precision Medicine. medRxiv 2024:2024.12.03.24318322. DOI: 10.1101/2024.12.03.24318322.

35. Kassirer JP. Our stubborn quest for diagnostic certainty. A cause of excessive testing. N Engl J Med 1989;320(22):1489–91. DOI: 10.1056/NEJM198906013202211.

36. Croskerry P. The cognitive imperative: thinking about how we think. Acad Emerg Med 2000;7(11):1223–31. (In eng). DOI: 10.1111/j.1553-2712.2000.tb00467.x.

37. Norman GR, Eva KW. Diagnostic error and clinical reasoning. Med Educ 2010;44(1):94-100. DOI: 10.1111/j.1365-2923.2009.03507.x.

38. Kassirer JP. Diagnostic reasoning. Ann Intern Med 1989;110(11):893-900. DOI: 10.7326/0003-4819-110-11-893.

39. Parikh RB, Teeple S, Navathe AS. Addressing Bias in Artificial Intelligence in Health Care. JAMA 2019;322(24):2377–2378. DOI: 10.1001/jama.2019.18058.

40. Char DS, Shah NH, Magnus D. Implementing Machine Learning in Health Care - Addressing Ethical Challenges. N Engl J Med 2018;378(11):981–983. DOI: 10.1056/NEJMp1714229.

41. Sabet C, Hammond A, Ravid N, Tong MS, Stanford FC. Harnessing big data for health equity through a comprehensive public database and data collection framework. NPJ Digit Med 2023;6(1):91. DOI: 10.1038/s41746-023-00844-5.

42. Goh E, Gallo R, Hom J, et al. Large Language Model Influence on Diagnostic Reasoning: A Randomized Clinical Trial. JAMA Netw Open 2024;7(10):e2440969. DOI: 10.1001/jamanetworkopen.2024.40969.

43. Torroba Hennigen L, Shen S, Nrusimha A, Gapp B, Sontag D, Kim Y. Towards Verifiable Text Generation with Symbolic References. (https://ui.adsabs.harvard.edu/abs/2023arXiv231109188T).

44. Li X, Zhu C, Li L, Yin Z, Sun T, Qiu X. LLatrieval: LLM-Verified Retrieval for Verifiable Generation. Mexico City, Mexico: Association for Computational Linguistics; 2024:5453-5471.

45. Chen S, Wong S, Chen L, Tian Y. Extending Context Window of Large Language Models via Positional Interpolation. (https://ui.adsabs.harvard.edu/abs/2023arXiv230615595C).

46. Wears RL, Perry SJ. Human factors and ergonomics in the emergency department. Ann Emerg Med 2002;40(2):206–12. DOI: 10.1067/mem.2002.124900.

47. Soltan AAS, Kouchaki S, Zhu T, et al. Rapid triage for COVID-19 using routine clinical data for patients attending hospital: development and prospective validation of an artificial intelligence screening test. Lancet Digit Health 2021;3(2):e78–e87. DOI: 10.1016/S2589-7500(20)30274-0.

48. Zegers M, Hesselink G, Geense W, Vincent C, Wollersheim H. Evidence-based interventions to reduce adverse events in hospitals: a systematic review of systematic reviews. BMJ Open 2016;6(9):e012555. (In eng). DOI: 10.1136/bmjopen- 2016-012555.

49. Li J, Lai Y, Li W, et al. Agent Hospital: A Simulacrum of Hospital with Evolvable Medical Agents. (https://ui.adsabs.harvard.edu/abs/2024arXiv240502957L).

50. Han S, Choi W. Development of a Large Language Model-based Multi-Agent Clinical Decision Support System for Korean Triage and Acuity Scale (KTAS)-Based Triage and Treatment Planning in Emergency Departments. (https://ui.adsabs.harvard.edu/abs/2024arXiv240807531H).

