## Supplemental Table S1 for "Grounded large language models for diagnostic prediction in real-world emergency department settings"

This material has been provided by the authors to give readers additional information about their work.

Table of contents

Supplementary Table S1: extracted parameters and definitions

### Supplementary Table S1: extracted parameters and definitions

| **Extracted parameter** | **Definition** |
| --- | --- |
| submission_ID | unique ID of the patient case submission |
| age | patient age at the time of consult, in years |
| sex | patient sex (m for male, f for female, u for unknown) |
| admission_time | timestamp of registration in the ED |
| main_complaint | main complaint, in a few words, as inputted by the ED admission secretary |
| heart_rate | heart rate (in beats per minute), at admission |
| systolic_BP | systolic blood pressure (in mmHg), at admission |
| diastolic_BP | diastolic blood pressure (in mmHg), at admission |
| saturation | peripheral oxygen blood saturation (in %), at admission |
| oxygen | external administration of oxygen? (Yes-no-unknown), at the time of saturation level registration |
| temperature | body temperature, in Celsius, at admission |
| GCS | Glasgow Coma Scale, to evaluate consciousness (on a 15 points scale), at admission |
| respiratory_rate | respiratory rate (in cycles/minute), at admission |
| past_medical_history | past medical history, physician-inputted |
| past_surgical_history | past surgical history, physician-inputted |
| treatment | usual medication taken, physician-inputted |
| treatment_verified | was the medication verified during ED visitation? (yes-no-unknown) |
| allergies | allergies |
| anamnesis | anamnesis, physician-inputted |
| physical_exam | physical examination |
